# Pregnant women as a sentinel population for genomic surveillance of malaria in the Democratic Republic of Congo

**DOI:** 10.1101/2024.05.27.24307472

**Authors:** Marie Onyamboko, Varanya Wasakul, Sarah Benie Bakomba, Daddy Kalala Kayembe, Bejos Kifakiou Nzambiwishe, Pascal Epe Ekombolo, Benjamen Basara Badjanga, Jean-Robert Moke Maindombe, Jephte Ndundu Ngavuka, Brunette Nsunda Lwadi, Eleanor Drury, Cristina Ariani, Sonia Goncalves, Vanapol Chamsukhee, Naomi Waithira, Tess D. Verschuuren, Sue J. Lee, Olivo Miotto, Caterina Fanello

## Abstract

Genomic surveillance is a valuable tool for detecting changes in the drug susceptibility of malaria parasites, allowing early modification of treatment strategies. However, implementation can be costly and problematic to set up in fragile and high-burden countries, especially when targeting cohorts of children. To address these challenges, we investigated whether in the Democratic Republic of Congo pregnant women attending antenatal care (ANC) services could act as a surrogate sentinel population. Between 2021 and 2023, we conducted a study in Kinshasa, targeting 4,001 pregnant women attending ANCs, and 2,794 children living in the same area. Blood samples from malaria-positive cases were genotyped using an amplicon sequencing platform, to allow comparisons of *Plasmodium falciparum* genomes between the two cohorts and estimations of drug-resistance mutations prevalence. Parasite populations sampled from the two cohorts exhibited highly similar allele frequencies at all tested loci, including drug resistance markers potentially under selection. Pregnant women did not have higher frequencies of sulphadoxine-pyrimethamine resistant haplotypes, which undermine preventive treatments, than children, and we did not find any *kelch13* mutation at significant frequency. Although parasite densities were lower in adults, the complexity of infection was similar to that in children. There was no evidence of *Plasmodium vivax* infections in the study. A cohort of pregnant women produces highly similar results to those from children, allowing the implementation of simple and efficient genomic surveillance systems integrated into routine ANC activities, while benefitting women with diagnosis and treatment. ClinicalTrials.gov Identifier: NCT05072613.

## Introduction

Drug resistance represents one of the major obstacles to the control and elimination of malaria. The emergence in sub-Saharan Africa of *Plasmodium falciparum* (*Pf*) parasites resistant to artemisinin (ART-R) is currently of particular concern, as this drug is the key component of artemisinin combination therapies (ACTs), currently used as first-line therapies in all countries in the region^1^. Large-scale genomic surveillance of *Pf* has been conducted for several years in Southeast Asia, where ART-R parasites have been circulating for more than a decade, and has proven a valuable tool for mapping resistant strains, detecting changes in antimalarial drug efficacy and informing public health authorities to allow rapid modification of preventive and curative treatment strategies^2,3^. The results of genomic surveillance complement those of more costly and complex *in vivo* therapeutic efficacy studies, which must be conducted in a more targeted manner, and are limited to observing the overall efficacy of an ACT. Conversely, genomic surveillance can simultaneously detect markers of resistance to individual ACT and of multiple other drugs, as well as changes in the dynamics of the parasite population, *e.g.*, in response to vaccine deployment. The Democratic Republic of Congo (DRC) is in the early stages of implementing genomic surveillance. The country remains one of the world’s most affected by the disease, with an estimated 31 million cases and 12% of global deaths attributable to malaria^4^. This epidemiological picture reflects the complexity of implementing control activities over a vast territory characterised by institutional and social fragility and ongoing conflict. In these settings, household and school surveys targeting children, the most affected group, present considerable challenges^5^. As a result, there is growing interest in leveraging on pregnant women attending antenatal care (ANC) services as a sentinel population for malaria surveillance, as they present fewer limitations than cohorts of children^6–12^. In areas of stable high malaria transmission, such as most of DRC, pregnant women are generally semi-immune to the clinical disease, and asymptomatic low-density infections are frequent. The lack of symptoms means that many episodes are not diagnosed, allowing infections to persist over time, with an increased risk of maternal anaemia and of adverse pregnancy outcome resulting from placental damage. Malaria testing at ANC level is beneficial to both mother and baby, and it also provides an opportunity to apply cost-effective sampling protocols for estimating malariometric parameters and to support surveillance.

In this study, we aimed to assess whether in an endemic area, pregnant women attending ANC services are a suitable sentinel population for genomic surveillance; in other words, whether samples collected from women can provide genomic surveillance results consistent with those collected in children from the same area. We present a detailed analysis of *Pf* genomes from samples collected in these two cohorts in Kinshasa, DRC capital, over a period of two years. In addition to a wide range of variants associated with drug resistance, our analysis compared allele frequencies at 100 loci thought not to be subject to selective pressures, to check for significant differences in the parasite populations. As an added benefit, the resulting data provides a report on the current levels of drug-resistance mutations, which we compared with those of studies conducted in previous years in Kinshasa.

## Methods

### Study design and population

This was a prospective study targeting pregnant women, regardless of their age or trimester of pregnancy and children under 14 living in the same area. Women were invited to participate and enrolled during their routine ANC visit. For children, we originally planned to conduct standard school-based surveys, but implementation was affected by the COVID-19 pandemic and subsequent vaccination campaign. We therefore adopted an alternative approach, setting up screening posts in existing health centres and, with the support of community health workers, encouraging families to visit the posts at their convenience. All study procedures were performed by trained research personnel. Each participant was included in the study once. The study was conducted in two areas of Kinshasa, urban and semirural, where malaria transmission is endemic and perennial^13^.

### Ethical aspects

The research adhered to the ethical principles outlined in the Declaration of Helsinki for medical studies involving human subjects. All participants (or their parents or guardians in the case of minors) provided written consent following a comprehensive explanation given in their preferred language prior to enrolment in the study. Approval was given by the research ethics committee of the University of Oxford (OxTREC, 548-21), the Kinshasa School of Public Health (ESP/CE133/2021) and the Ministry of Health of DRC (025/GPK/CAB/MIN.SPH/LNK/NIP/ski/2021). Further approval was granted for screening school children by the Provincial Ministry of Health, the Provincial Ministry of Education, the Provincial Division Head and the National Intelligence Agency. Approval for exporting blood samples was granted by the Ministry of the Environment and Sustainable Development in accordance with the Nagoya Protocol. ClinicalTrials.gov Identifier: NCT05072613.

### Samples and data collection

Participants were tested for malaria with a *pfhrp2*-based rapid diagnostic test (RDT, SD Bioline™ Malaria Ag P.f/Pan) and haemoglobin levels were measured with HemoCue^®^ 301 (Angelholm, Sweden). A capillary blood sample was taken from each RDT-positive participant, to determine the parasite species and density by standard microscopy, and to prepare a dried blood spot (DBS) for genomic analysis. Individuals with malaria and/or anaemia were offered treatment as per DRC national guidelines or referred to hospital if severe. Demographic and anthropometric data were collected from all participants. Epidemiological data were entered locally using the REDCap electronic data capture tool hosted by the University of Oxford.

### Statistical analysis

A sample size of 650 DBSs from each cohort was estimated to enable accurate estimation of the prevalence (up to 50% +/-4%) of antimalarial drug resistance associated mutations with 95% confidence. Since malaria prevalence in this area ranges from 15 to 30%^14^, we aimed to screen a minimum of 3,540 individuals in each cohort over a 12-month period. Comparisons used the chi-squared or Kruskal-Wallis test, and correlations were assessed using Kendall’s tau. A two-proportion z-test was used to compare allele frequencies; the 95% confidence interval for proportion was calculated by Agresti-Coull method (BinomCI; DescTools R package version 0.99.54^15^). Logistic regression was used to account for confounders; in particular, since the proportions of women and children differed between urban and semirural areas (Table 1), we adjusted for area when comparing allele frequencies. Parasite levels were log-transformed for the analysis. Data were analysed using STATA version 18.0 (StataCorp LLC, USA) and R version 4.2.3.

**Table 1.** Summary of demographic, anthropometric and laboratory results in the two cohorts (children vs pregnant women)

| Characteristics | Children | Pregnant women |
| --- | --- | --- |
| **n Enrolled** | 2,794 | 4,001 |
| Schools, n (%) | 92 (3.3) | - |
| Health Centres, n (%) | 2,702 (96.7) | - |
| Antenatal Care Service, n (%) | - | 4,001 (100) |
| Urban, n (%) | 892 (22.6) | 3057 (77.4) |
| Semirural, n (%) | 1902 (66.8) | 944 (33.2) |
| Female, n (%) | 1,388 (49.7) | - |
| Median age (IQR), years | 6.1 (3.0, 9.4) | 28.7 (24.0, 33.4) |
| Date of birth unknown, n (%) | 18 (0.6) | 39 (1.0) |
| Pregnant teenagers (<19 y.o.), n (%) | - | 220 (5.5) |
| Primiparae, n (%) | - | 758 (19.0) |
| Median body temperature (IQR), °C | 36.5 (36.2, 36.8) | 36.3 (36.2, 36.6) |
| Fever (>37.5 °C), n (%) | 215 (7.7) | 7 (0.2) |
| Report of sickness in the previous days, n (%) | 1,989 (71.2) | - |
| *P. falciparum* by RDT, n (%) | 1,368 (49.0) | 765 (19.1) |
| *P. falciparum* by microscopy, n (%) | 1,180 (86.3) | 703 (91.9) |
| Geo. mean parasitaemia/μL (95% CI) | 5,998 (5,031 to 7,150) | 571 (486 to 672) |
| Hyperparasitaemia (>100,000/μL), n (%) | 236 (20.0) | 3 (0.4) |
| Dried Blood Spots with available results | 1,356 | 644 |

### Genomic analysis

DBSs were processed by the Wellcome Sanger Institute (WSI; UK), using the SpotMalaria *Plasmodium* genotyping platform^2^. Data were encoded into Genetic Report Card format, comprising genotypes at drug resistance-related single-nucleotide polymorphisms (SNPs), as well as genetic barcodes consisting of genotypes at 100 biallelic SNPs known to be variable globally, and not related to drug resistance^2^. In any given sample, SNPs with insufficient read coverage to call a genotype were encoded as *missing*, while SNPs with reads for both alleles were called as *heterozygous*. Sample *missingness* (a measure of genotyping quality) was estimated as the proportion of barcoding SNPs called as *missing*, while sample *heterozygosity* was estimated as the proportion of non-missing barcoding SNPs called as heterozygous. Samples with missingness >50% were deemed of low genotyping quality and excluded from heterozygosity analyses. At any given SNP, the *non-reference allele frequency* (NRAF) in a population was determined by excluding samples with missing and heterozygous calls, and then determining the proportion of the remaining samples that carried the *non-reference* allele. Infecting *Plasmodium* species were identified by kraken2 (version 2.0.7)^16^ and summarized with krakentools (version 1.2), using a custom database comprising mitochondrial sequences of six *Plasmodium* species (Supplementary Table 1). We deemed a species to be present if the classifier assigned >=5 reads and >=1% of reads for that sample. To analyze changes in drug resistant allele frequencies over time, we compared the frequencies in the cohorts of children with those of studies conducted between 2012 and 2016 in Kinshasa by the same research team and available from the MalariaGEN Pf7 dataset^17^.

## Results

### Summary of epidemiological features of malaria

Between November 2021 and June 2023, 2,794 children and 4,001 pregnant women were recruited; a summary of demographic, anthropometric, and laboratory results by cohort is presented in Table 1. Malaria prevalence by RDT was 49·0% (95% CI 47·1-50·8) in children and 19·1% (95% CI 17·9-20·3) in women. About 14% of RDT-positive cases among children and 8% among women were negative by microscopy, with the larger discrepancy observed in the first group (p<0·001) likely to result from a higher number of recently treated infections with detectable *Pf*HRP2 antigen levels. Within each cohort, parasite densities correlated negatively with age (Kendall’s Tau: -0·057, p=0·003 in children; - 0·159, p<0·001 in women). Women had comparable prevalences of malaria infections in urban and semirural area (p=0·228), while parasite levels were higher in semirural area (p<0·001; Supplementary Table 2), most likely because participants were on average younger and more primiparous. Children living in semirural area were significantly more infected and with higher parasite levels than those living in the urban area (p<0·001, Supplementary Table 2), a difference that may be partly attributed to the free routine malaria screening and treatment provided by our center (located in the semirural area), which attracts more sick children. The final sample size was larger than planned, to account for the higher prevalence of malaria in the child cohort, the need to sample both cohorts simultaneously, and the discrepancy observed between microscopy and RDT results.

### *Plasmodium* species identification

Genomic results were returned from WSI for samples from 644 pregnant women and 1,356 children. We could identify the infecting species in 1,967 of the 2,000 samples analysed (98·3%), all of which contained *Pf* parasites. Co-infections with *P. malariae* were detected in 160 samples (8·1%), and with *P. ovale* in 56 (2·8%); both species were found in varying proportions within samples (1-99% of sequencing reads). We found no evidence of other *Plasmodium* species, including *P. vivax* and *P. knowlesi*.

### Barcoding allele frequencies comparison

To verify whether allele frequencies in samples from pregnant women deviated significantly from those in children, we estimated NRAF in the two cohorts at each of 100 barcoding SNPs, which were chosen for their variability at a global level but are not thought to be under selection^2^. We found an excellent correlation between the NRAF in the two cohorts (Figure 1; Pearson correlation coefficient r=0·99, p<0·001). Only two variants showed statistically significant differences between cohorts (barcoding SNPs 95 and 37; p<0·01), but in both cases the differences were moderate: 0·22 [95% CI 0·18-0·26] *vs* 0·14 [0·12-0·16] at SNP 95, and 0·40 [0·33-0·43] *vs* 0·29 [0·26-0·33] at SNP 37 in women and children, respectively (Supplementary Figure 1). At all remaining SNPs, confidence intervals of the frequency estimates overlapped, indicating that parasite populations were minimally differentiated between cohorts.

**Figure 1.**
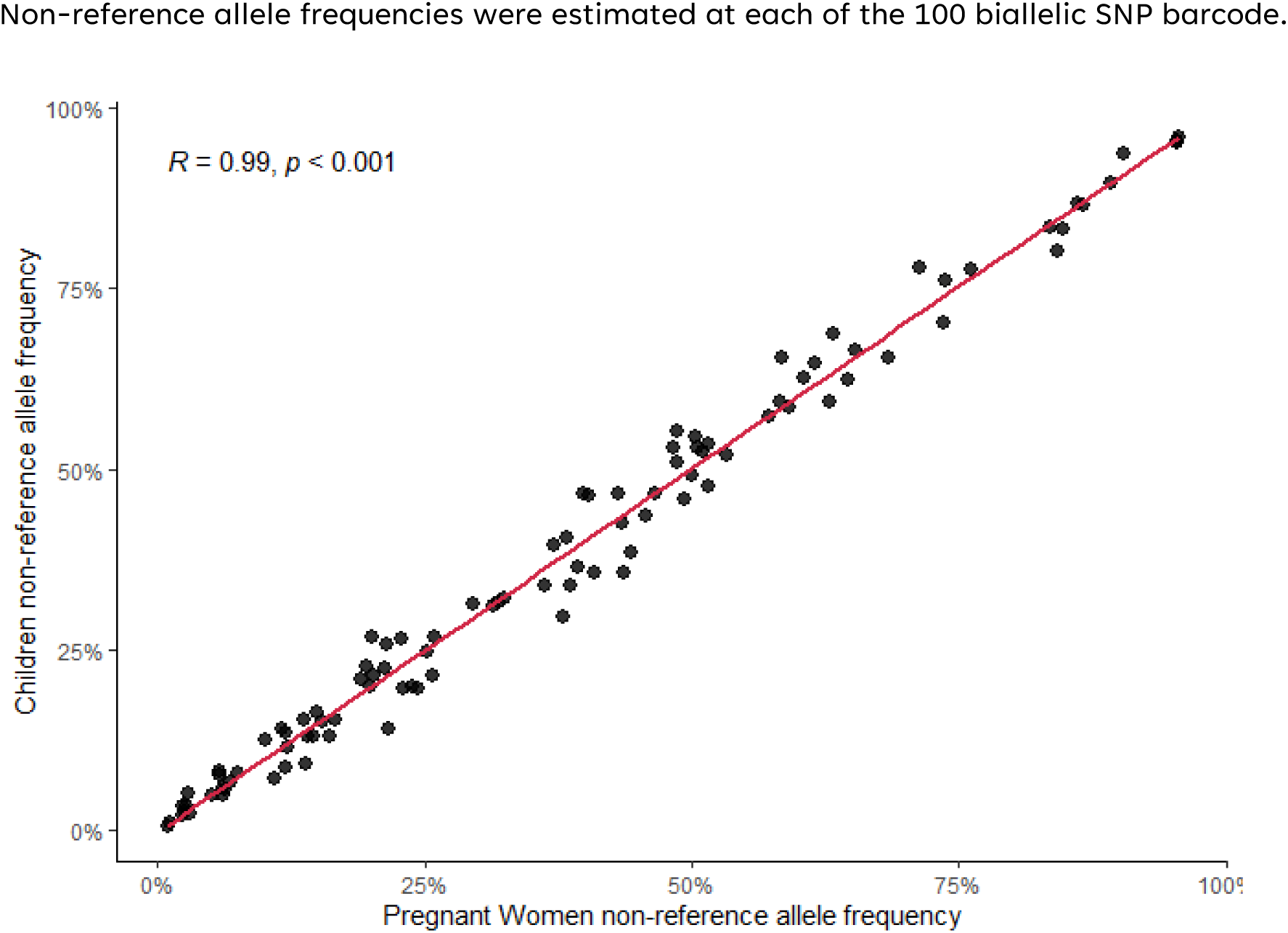
Non-reference allele frequency correlation between cohorts Non-reference allele frequencies were estimated at each of the 100 biallelic SNP barcode.

### Frequencies of antimalarial resistance markers

We also estimated allele frequencies at a number of known drug resistance loci that might be under selection pressure in DRC (Table 2). Allele differences at these loci would suggest that parasites infecting the two cohorts are subjected to different selective pressures. We were able to fully scan the kelch and BTB/POZ domains of the *kelch13* gene (PF3D7_1343700)^18^ in 1,316 samples, but did not find any mutation associated with ART-R at elevated frequency. Besides the A578S variant, which has been shown not to confer ART-R^19^, only the V534A mutation, observed in two cases, was found as a homozygous variant. Six other mutations (P527H, S522C, G533S, Q613E, I634L and D641N) were each found in a single mixed infection. The low frequency of these mutations, and their presence in mixed infections, are consistent with expected prevalence of these variants in African countries under normal conditions^20^.

**Table 2.** Comparison of allele frequencies in the two cohorts at SNPs associated with antimalarial drug resistance. This table shows allele frequencies at positions that were tested for known mutations associated with drug resistance. Each row represents a SNP, with columns showing: the chromosome and position of the SNP, the reference and alternative (Nref) alleles; the frequency of the alternative allele in the two cohorts; the p-value of logistic regression adjusted by area to determine whether the mutation prevalence is significantly different in the two cohorts-significant values (p<0·01) are shown in red. The last column indicated, for *dhfr* and *dhps* SNPs only, the sulfadoxine-pyrimethamine (SP) resistant haplotype that includes the mutation.

| Gene | Position | Ref | Nref | Non-reference Allele Frequencies |  | p-value | SP Haplotype |
| --- | --- | --- | --- | --- | --- | --- | --- |
|  |  |  |  | Children | Pregnant Women |  |  |
| *dhfr* | 51 | N | I | 1107 / 1113 (99.5%) | 499 / 503 (99.2%) | 0.842 | Triple |
| *dhfr* | 59 | C | R | 932 / 1004 (92.8%) | 430 / 463 (92.9%) | 0.816 | Triple |
| *dhfr* | 108 | S | N | 1097 / 1103 (99.5%) | 475 / 478 (99.4%) | 0.664 | Triple |
| *dhfr* | 164 | I | L | 0 / 1101 (0.0%) | 0 / 479 (0.0%) | 1.000 | Sextuple |
| *dhps* | 436 | S | A | 74 / 1014 (7.3%) | 40 / 433 (9.2%) | 0.021 |  |
| *dhps* | 437 | A | G | 1038 / 1067 (97.3%) | 435 / 444 (98.0%) | 0.456 | Quintuple |
| *dhps* | 540 | K | E | 185 / 889 (20.8%) | 59 / 339 (17.4%) | 0.922 | Quintuple |
| *dhps* | 581 | A | G | 112 / 921 (12.2%) | 40 / 348 (11.5%) | 0.566 | Sextuple |
| *dhps* | 613 | A | S | 34 / 993 (3.4%) | 17 / 373 (4.6%) | 0.035 | Sextuple |
| *crt* | 74 | M | I | 131 / 1016 (12.9%) | 36 / 448 (8.0%) | 0.110 |  |
| *crt* | 75 | N | E | 129 / 1011 (12.8%) | 37 / 447 (8.3%) | 0.145 |  |
| *crt* | 76 | K | T | 132 / 1016 (13.0%) | 39 / 453 (8.6%) | 0.160 |  |
| *crt* | 220 | A | S | 157 / 959 (16.4%) | 49 / 456 (10.7%) | 0.069 |  |
| *crt* | 271 | Q | E | 143 / 969 (14.8%) | 41 / 451 (9.1%) | 0.075 |  |
| *crt* | 356 | T | I | 104 / 1009 (10.3%) | 40 / 450 (8.9%) | 0.891 |  |
| *crt* | 371 | R | I | 156 / 976 (16.0%) | 49 / 465 (10.5%) | 0.155 |  |
| *mdr1* | 86 | N | Y | 54 / 1030 (5.2%) | 27 / 431 (6.3%) | 0.230 |  |
| *mdr1* | 184 | Y | F | 226 / 798 (28.3%) | 103 / 350 (29.4%) | 0.214 |  |
| *mdr1* | 1246 | D | Y | 5 / 1011 (0.5%) | 0 / 334 (0.0%) | 0.992 |  |

We found minimal or non-significant differences between cohorts in variants within the *dhfr* (PF3D7_0417200) and *dhps* (PF3D7_0810800) genes associated with antifolate resistance (Table 2). This is noteworthy, since sulfadoxine-pyrimethamine (SP) is used for chemoprevention in pregnancy (Intermittent Preventive Treatment in pregnancy, IPTp-SP) and could potentially exert selective pressure on parasites infecting pregnant women. We observed several combinations of *dhfr* and *dhps* mutations that have been associated with reduced parasite sensitivity to SP. The *triple mutant* haplotype (comprising *dhfr* N51I, C59R and S108N), which confers resistance to pyrimethamine^21^ was found in 92·5% of the isolates (Table 3) while the *quintuple mutant* haplotype (triple mutant with additional mutations A437G and K540E in *dhps*) associated with failure to SP^21^ was detected in 20·2% of the isolates. The sextuple mutant haplotype (carrying the additional *dhps* A581G mutation) that further reduces SP sensitivity and compromises IPTp-SP efficacy^21^, was detected in 9·2% of samples.

**Table 3.**
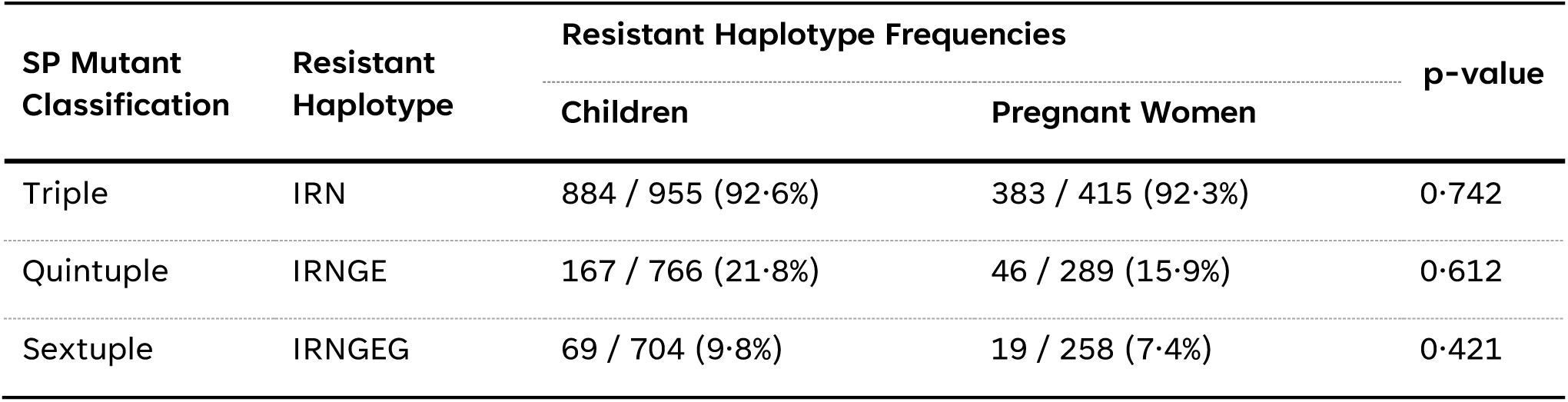
Comparison of sulphadoxine-pyrimethamine resistant haplotypes in the two cohorts. This table shows the frequencies of haplotypes resistant to sulfadoxine-pyrimethamine (SP). Each row represents a haplotype, while the columns show: classification and associated haplotype; frequency of the resistant haplotype for each cohort; the p-value of the logistic regression to determine whether the prevalence of the haplotype is significantly different in the two cohorts. The triple mutant contains the *dhfr* mutations N51I, C59R, S108N; the quintuple mutant contains the *dhfr* triple and two additional *dhps* mutations (A437G, K540E); the sextuple mutant has an additional mutation at *dhfr* I164L or *dhps* A581G or *dhps* A613S/T. Only one form of the sextuple mutant (IRNGEG) was identified in the present study.

Furthermore, the *dhps* A581G mutation was also detected in isolation (*i.e.,* not as part of a sextuple haplotype) in about one-third of samples; similarly, the *dhps* A613S mutation, an alternative variant of the sextuple haplotype, was detected in isolation in 43 samples. The *dhfr* I164L mutation, also a potential sextuple mutant component, was absent in our dataset, consistent with its low prevalence in Africa^22^.

The analysis of polymorphisms in the *crt* gene (PF3D7_0709000) associated with resistance to chloroquine and amodiaquine^23,24^ also did not reveal significant differences between cohorts. The most common chloroquine-resistant haplotype (often referred to as ‘CVIET’ for its amino acids at positions 72-76) was detected in 12·8% in children and 8·1% in pregnant women (p=0·128 adjusted by area). The *crt* gene mutations associated with piperaquine resistance in Southeast Asia (*e.g.*, at positions 97, 218 and 333) were absent from our sample set.

Mutations in the *mdr1* gene (PF3D7_0523000) were generally at low levels, with the exception of the Y184F mutation, which was present in 28·7% of samples, without significant differences between cohorts.

### Temporal trends of antimalarial resistance markers

A comparison of current allele frequencies at drug-resistant loci in *crt*, *dhps, dhfr* and *mdr1*, against 573 samples collected in 2012-16, showed major changes in drug resistance profiles (Figure 2). The frequency of the CVIET haplotype decreased significantly: from 61·6% in 2012-16 to 12·8% in 2021-23 (p<0·001). Similarly, the mutation at position N86Y on *mdr1* decreased significantly from 32·2% to 5·2% (p<0·001). In contrast, no differences were found in the frequency of the Y184F mutation. This analysis showed a substantial increase in the prevalence of all haplotypes conferring resistance to SP, largely driven by a significant increase in the frequencies of the *dhps* K540E and A581G mutations (540E from 7·9% to 20·8%, p<0·001; 581G from 3·0% to 12·2%, p <0·001).

**Figure 2.**
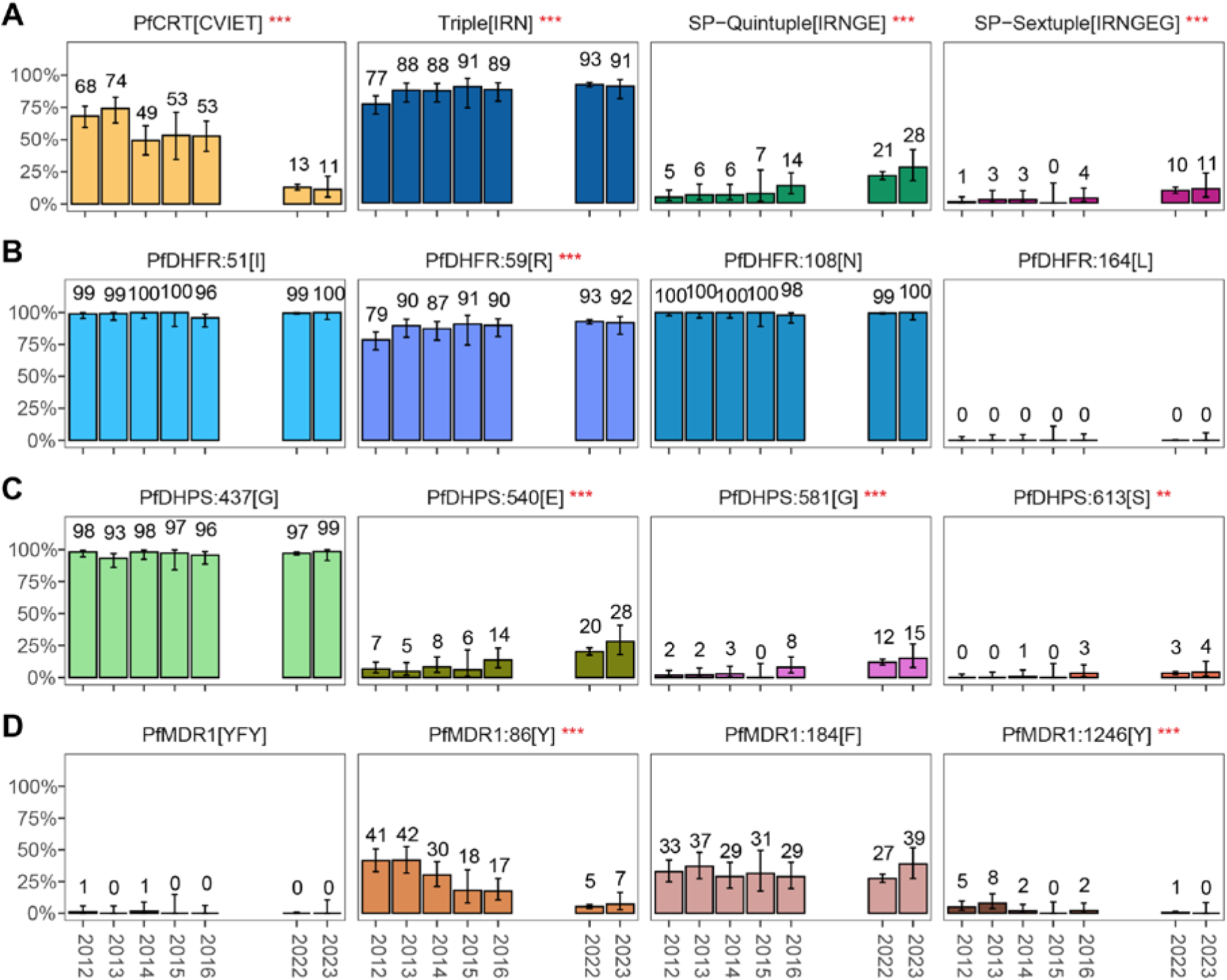
Temporal trends of drug-resistant haplotypes and alleles in children from Kinshasa. The values represent yearly proportion of A) haplotypes conferring chloroquine (PfCRT) and sulphadoxine-pyrimethamine resistance (triple, quintuple and sextuple), B) dhfr alleles, C) dhps alleles D) and mdr1 haplotype and alleles. Resistant haplotypes or amino acids are indicated in square brackets. Data from 2012 to 2016 are from Pf7 data resource, while data from 2022 to 2023 are from the present study. Fifteen samples, collected towards the end of 2021 were aggregated with data from 2022. Error bars indicate 95% confidence intervals. Two-proportion z-tests were conducted to compare the period 2012–2016 with that of 2022–2023. Significant differences between the two periods are denoted above the plot as follows: ***p<0·001, **p<0·01, *p<0·05.

### Genotype missingness

Elevated genotype missingness, typically caused by low parasite DNA concentration in the sample (*e.g.,* because of low parasite density) can adversely affect genomic analyses, especially those in which incomplete haplotypes must be excluded. In this study, we observed a strong inverse correlation between parasite densities and genotype missingness in both cohorts (Supplementary Figure 2; Kendall’s Tau = -0·437 and -0·510 in women and children respectively, both p<0·001). This suggests that applying a minimum parasitaemia threshold may increase cost-efficiency of surveillance, and also that ANC-population might produce slightly smaller sample sizes for certain types of analyses.

### Complexity of infection

We used the *heterozygosity level*, defined here as the proportion of non-missing barcode SNPs assigned a heterozygous genotype, as a measure of the *complexity of infection* (COI). Heterozygosity levels correlate well with COI estimates predicted by the RealMcCoil bioinformatic tool^25^. COI estimates ranged from 1 to 5, with approximately 50% of infections being polyclonal (COI ≥2) in both cohorts. The median (IQR) heterozygosity level was 0·16 (0·01, 0·35), and although children had slightly higher values than women [0·17 (0·10, 0·37) *vs* 0·14 (0·02, 0·30), p=0·15] Supplementary Figure 3), age and heterozygosity were not correlated (Kendall’s Tau =-0·015; p=0·368).

## Discussion

Genomic surveillance has proven itself a very useful tool for informing malaria control programmes tracking the emergence and spread of antimalarial drug resistant strains. Yet, the most fragile countries, which are also those with the highest burden of disease and thus could benefit most from the availability of new tools, are also those with the greatest difficulties in implementation. There is an imperative to find strategies that can simplify genomic surveillance implementations and integrate them seamlessly in public health processes. This is particularly important for malaria, where the people most at risk are also the most vulnerable and therefore most problematic to monitor. In this work, we made important steps towards overcoming some key implementation issues: we have established that a cohort of adults, passively screened at ANC facilities, will produce the same genomic surveillance results as a more difficult to monitor cohort of children. In most DRC, as many other African countries, ANC services are available throughout the territory, and women visit these facilities at least once during pregnancy. Leveraging on ANCs can lead not only to simpler and more economical implementations of surveillance, but also to the detection and treatment of malaria in asymptomatic women and complement other prevention strategies in pregnancy where uptake is low.

An important finding from this study is that allele frequencies estimated from women were very similar to those from children, not only at neutral barcoding SNPs, but also for mutations associated with resistance to antimalarial drugs. This was by no means a given: since women and children are not exposed to the same antimalarial drugs, and exhibit different age-related responses to infection, differences could not be ruled out. Notably, the two parasite populations had several other similar characteristics. For example, levels of complexity of infection, which result from bites of mosquitoes infected with more than one clone and/or multiple bites, were similar in both cohorts, even though women tend to harbor longer-lasting asymptomatic parasite reservoirs. These pronounced levels of heterozygosity suggest that pregnant women can also provide valuable information about changes in transmission intensity^26^, *e.g*., in response to vaccine deployment.

In the process of demonstrating the equivalence of the two cohorts, we have also estimated current levels of drug resistant mutations prevalence in Kinshasa and described their historic trends for the last decade. Reassuringly, we found no evidence of ART-R mutations in *kelch13*, which are increasingly being reported in East Africa^1^ and, as it now appears, the eastern borders of DRC^27^; although geographically distant from the areas affected, Kinshasa is a recipient for internally displaced people from other provinces, particularly from eastern DRC, and could become a spreading hub. Resistance to antifolates, namely SP, poses a more pressing problem, because of widespread highly resistant haplotypes in the *dhfr* and *dhps* genes, which undermine preventative treatments. In Kinshasa we found more than 90% of infections harbouring triple mutant haplotypes, 20% carrying quintuple, fully resistant haplotypes, and about 10% with the sextuple super-resistant haplotype.

Interestingly, two mutations normally considered part of sextuple mutant haplotypes (*dhps* A613S/T and A581G) were frequently found without the remainder of the haplotypes, highlighting the variability of the *dhps* gene^28,29^. The frequency of resistant haplotypes, and the evidence of an increasing trend in some mutations over time, suggest continued pharmacological pressure from SP, which is mainly used in DRC for intermittent preventive treatment during pregnancy. Although up to a certain level of resistance the benefits of IPTp-SP may still outweigh the risks of adverse birth outcomes, our surveillance results suggest that the country should start evaluating chemopreventive alternatives, such as dihydroartemisinin-piperaquine, which has been shown to significantly improve maternal health compared to SP^30^. Analyses of historical trends also showed that resistance to the 4-aminoquinoline chloroquine, mediated by mutations in the *crt* gene, was low with evidence of a temporal decline over the past decade. This downward trend was accompanied by a decline in the frequency of the N86Y mutation on the *mdr1* gene; the role of this mutation is unclear, as it has been associated with reduced sensitivity to amodiaquine (another 4-aminoquinoline) but there is some evidence that lumefantrine may select for the wild-type allele^31^. Since in DRC amodiaquine-artesunate and artemether-lumefantrine are both part of a first-line multitherapy strategy, and both currently available on the market, the *mdr1* N86Y trend remains difficult to interpret. Reliable, validated genetic markers for resistance to amodiaquine and lumefantrine are urgently needed, if critically-needed monitoring of ACT partner drug efficacy in Africa is to be supported.

It is important to note that the use of ANC sentinel population has some drawbacks that need to be accounted for. Although about 20% of women were found to be infected, providing an abundant supply of samples, their lower parasite densities translate to lower parasite DNA content, and therefore a higher chance of genotyping failure. To avoid wastage, genomic surveillance studies could establish a minimum parasitaemia threshold, and screen samples accordingly. Arguably, this would also be a problem if sampling from school-children, given that they also tend to have low parasitaemia densities, partly because sick children would not be present on the day of the survey, and also because disadvantaged children, who are most at risk of malaria, have low school attendance.

Pragmatic implementation strategies, such as the one we recommend here, may only be enabling components of more complex implementations. There are numerous other systemic obstacles to implementing genomic surveillance in low-resource settings, such as difficulties in building and maintaining local technical capacity and the costs and delays associated with procurement of consumables. Suitable strategic choices will only succeed with donor support, alignment with ongoing capacity-building efforts in Africa, and input from the international scientific community. The benefits are substantial, as surveillance can be applied to other pathogens than malaria as well as to antimicrobial resistance.

## Author’s contributions

Study design, data collection, data analysis, manuscript preparation: CF, OM, SJL, VW, NM, TV, MO. Data collection: SBK, DKK, BKN, PEE. Laboratory analysis: BBB, JMM, JNN, BNL, ED, CA, SG. Data management: NW, VC. For the purpose of open access, the authors have applied a CC BY public copyright license to any Author Accepted Manuscript version arising from this submission.

## Funding

This study was funded by Bill & Melinda Gates Foundation (grant numbers INV-033243 and INV-007590) and the Wellcome Trust (220540/Z/20/A).

## Data availability

Final genomic data will be made available in a public repository upon publication, and in the interim will available by contacting the corresponding author. De-identified participant data will be available from the Mahidol Oxford Tropical Medicine Data Access Committee upon request from this link: https://www.tropmedres.ac/units/moru-bangkok/bioethics-engagement/datasharing.

## Competing interests

None to be declared.

## Acknowledgments

We thank all women and children who participated in our study, the staff of KIMORU, and the staff and management of the participating ANCs and Health Centres for hosting the study. Genome sequencing and genotyping was performed by the Wellcome Sanger Institute (WSI), and sequencing data processing was supported by the MalariaGEN Resource Centre. We thank the staff of the WSI Sample Logistics, Sequencing, and Informatics facilities for their contribution and Victoria Simpson for coordinating the MalariaGEN Resource Centre. This publication uses data from the MalariaGEN *Plasmodium falciparum* Community Project as described in https://doi.org/10.12688/wellcomeopenres.16168.2.

## Supplementary Materials

**Supplementary Table 1.**
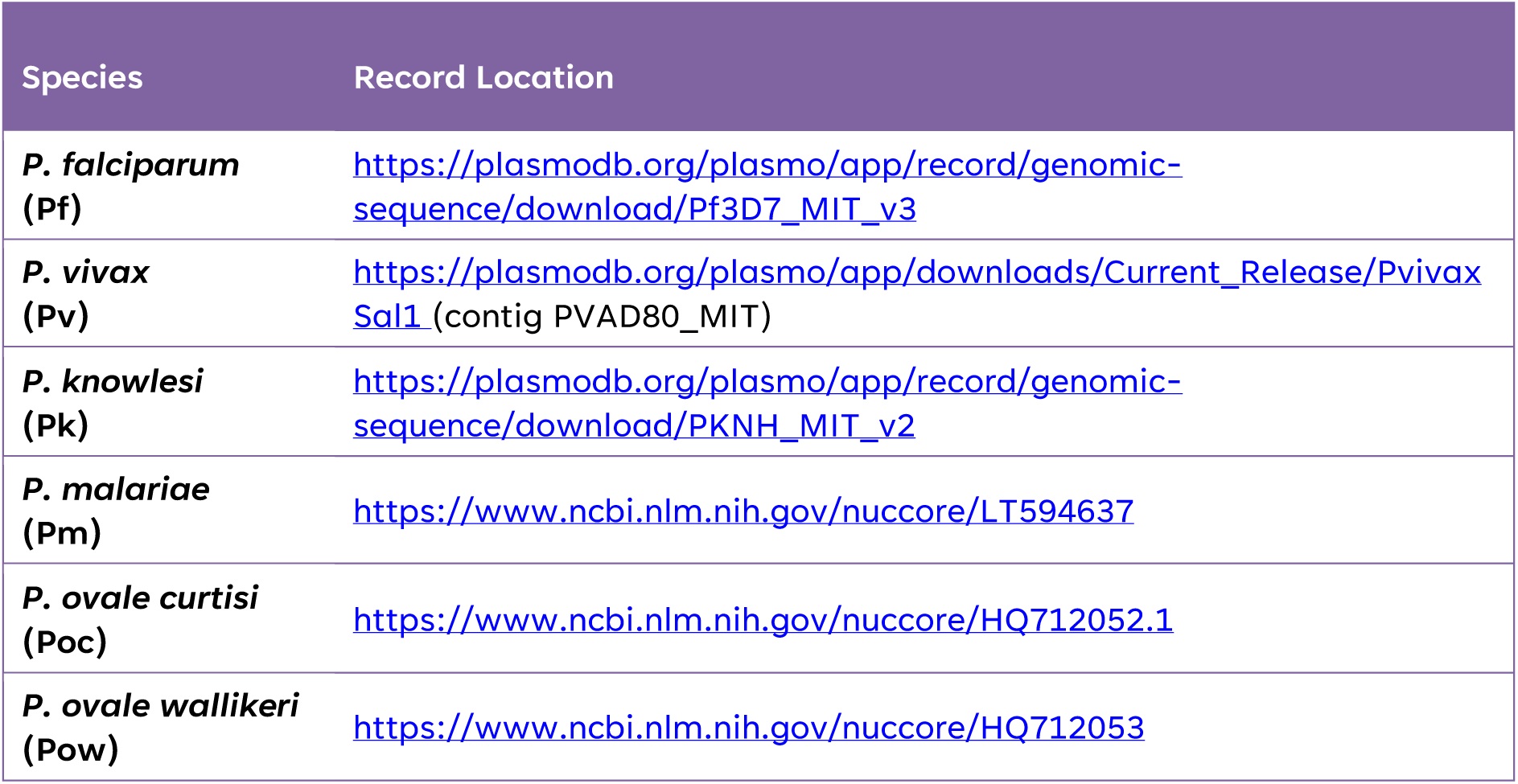
Location of mitochondrial sequence sources used to classify six *Plasmodium* species.

| Species | Record Location |
| --- | --- |
| *P. falciparum* (Pf) | https://plasmodb.org/plasmo/app/record/genomic-sequence/download/Pf3D7_MIT_v3 |
| *P. vivax* (Pv) | https://plasmodb.org/plasmo/app/downloads/Current_Release/Pvivax_Sal1 (contig PVAD80_MIT) |
| *P. knowlesi* (Pk) | https://plasmodb.org/plasmo/app/record/genomic-sequence/download/PKNH_MIT_v2 |
| *P. malariae* (Pm) | https://www.ncbi.nlm.nih.gov/nuccore/LT594637 |
| *P. ovale curtisi* (Poc) | https://www.ncbi.nlm.nih.gov/nuccore/HQ712052.1 |
| *P. ovale wallikeri* (Pow) | https://www.ncbi.nlm.nih.gov/nuccore/HQ712053 |

**Supplementary Table 2.** Summary of *P. falciparum* characteristics in the two cohorts by area.

| Cohort | Urban | Semirural | p-value |
| --- | --- | --- | --- |
| **Women** |  |  |  |
| *P. falciparum* by RDT, N (%) | 598 (19.6) | 167 (17.8) | 0.228 |
| Positive cases by microscopy, N (%) | 548 (92.8) | 155 (91.6) | 0.623 |
| Geo. mean parasitaemia/μL (95% CI) | 502 (417-605) | 889 (647-1,221) | <0.001 |
| **Children** |  |  |  |
| *P. falciparum* by RDT, N (%) | 318 (55.2) | 1,050 (35.7) | <0.001 |
| Positive cases by microscopy, N (%) | 275 (86.5) | 905 (86.2) | 0.896 |
| Geo. mean parasitaemia/μL (95% CI) | 2,361 (1,641-3,397) | 7,961 (6,535-9,699) | <0.001 |

**Supplementary Figure 1.**
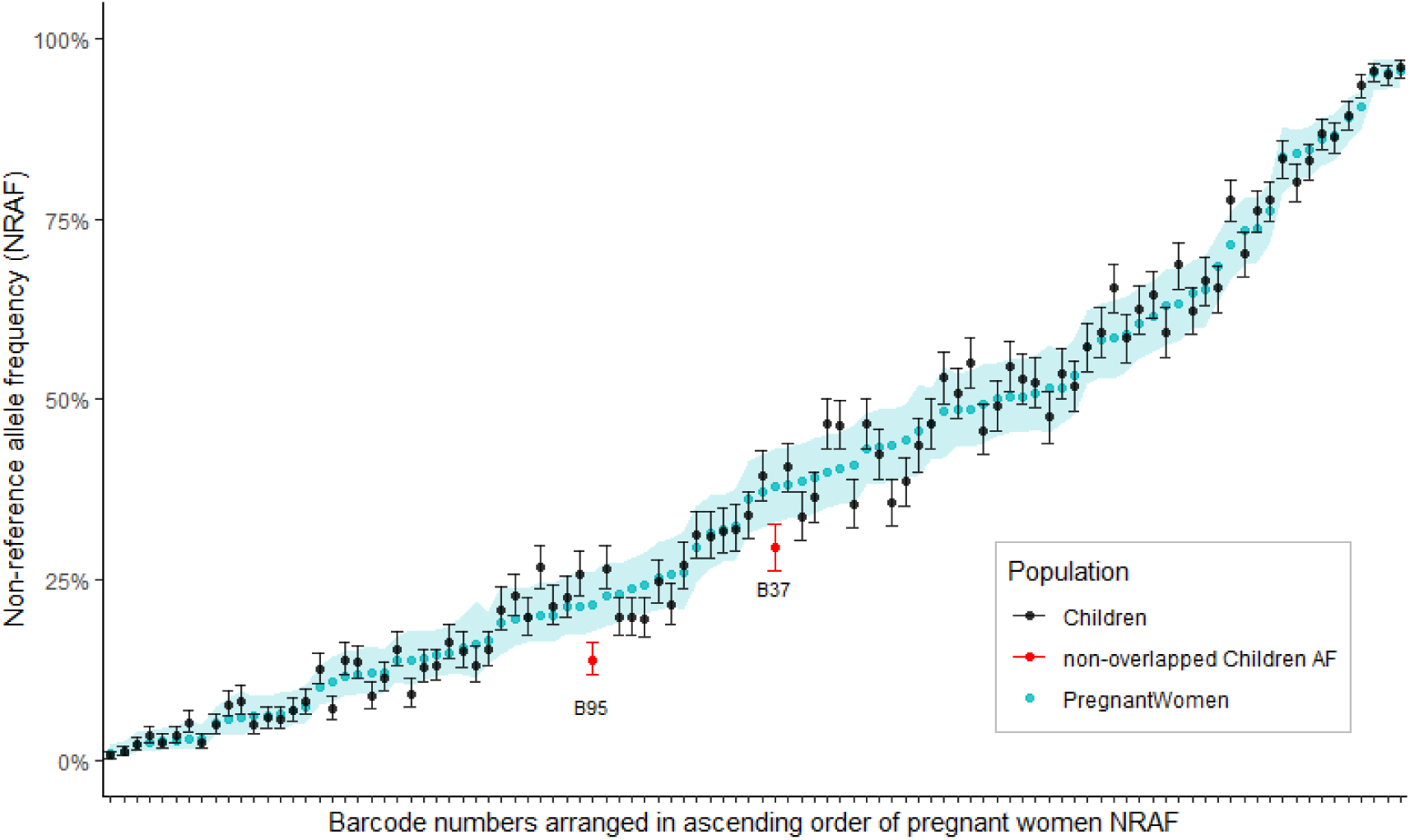
Estimated non-reference allele frequencies (NRAF) and 95% confidence intervals at each barcode SNP position in each cohort. Overlapping of confidence intervals of the frequency estimates indicates minimal differentiation in the population between the two cohorts. Pregnant women’s NRAF are coloured in blue with 95% confidence intervals shaded in. Children’s NRAF are in black with error bars indicating 95% confidence interval. Red colour highlights children’s NRAF where there are non-overlapping confidence intervals between the two cohorts (Barcode SNP #95 and #37). To improve visualisation, barcode SNP positions are arranged in ascending order of pregnant women’s NRAF.

**Supplementary Figure 2.**
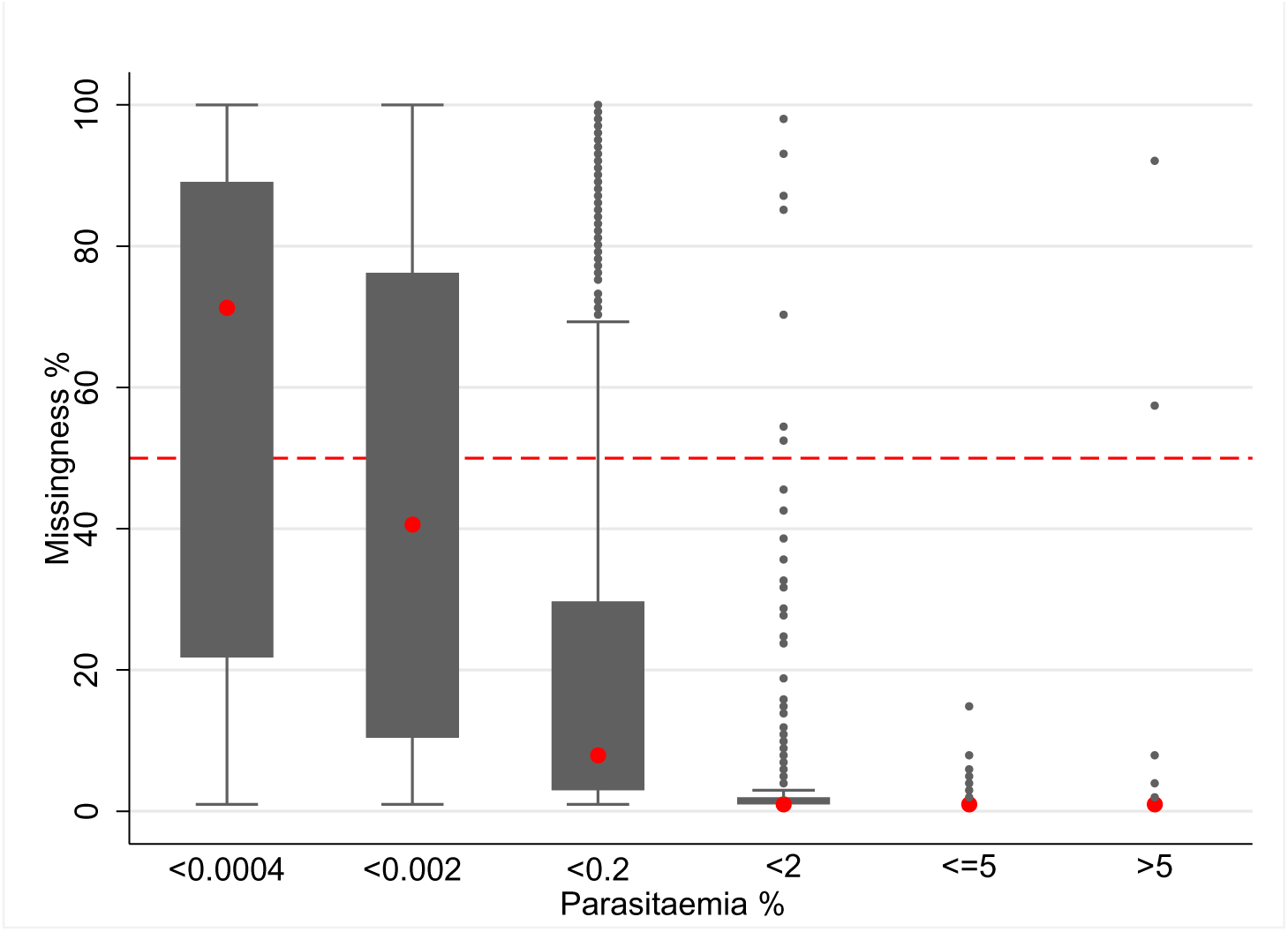
Relationship between parasitaemia and genotype missingness. Values on the y-axis represent the percentage missingness and values on the x-axis the percentage parasitaemia. Each box represents the median and interquartile range of percentage parasitaemia by microscopy. The red dash line indicates 50% level of missingness.

**Supplementary Figure 3.**
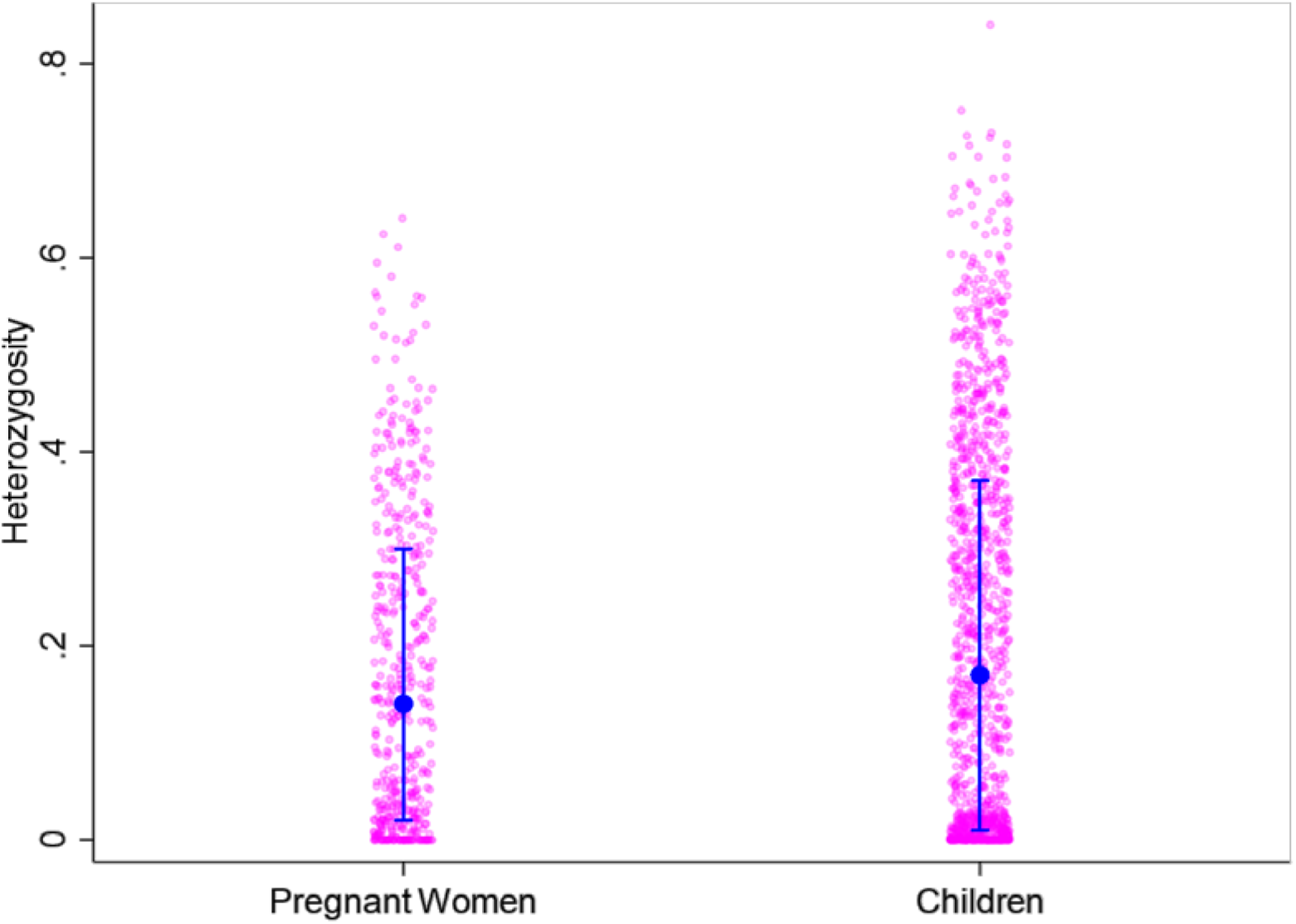
Heterozygosity in the two cohorts. Each pink dot represents the percentage heterozygosity for one individual sample; each capped bar represents a population (pregnant women and children) with 95% percent Confidence Intervals.

## Notes

### Competing Interest Statement

The authors have declared no competing interest.

### Funding Statement

This study was funded by Bill & Melinda Gates Foundation and the Wellcome Trust of England.

### Author Declarations

Approval was given by the research ethics committee of the University of Oxford (OxTREC, 548-21), the Kinshasa School of Public Health (ESP/CE133/2021) and the Ministry of Health of DRC (025/GPK/CAB/MIN.SPH/LNK/NIP/ski/2021).

## References

1. Rosenthal PJ, Asua V, Conrad MD. Emergence, transmission dynamics and mechanisms of artemisinin partial resistance in malaria parasites in Africa. Nat Rev Microbiol 2024.

2. Jacob CG, Thuy-Nhien N, Mayxay M, et al. Genetic surveillance in the Greater Mekong subregion and South Asia to support malaria control and elimination. Elife 2021; 10.

3. Noviyanti R, Miotto O, Barry A, et al. Implementing parasite genotyping into national surveillance frameworks: feedback from control programmes and researchers in the Asia-Pacific region. Malar J 2020; 19(1): 271.

4. WHO. World Malaria Report: World Health Organization; 2023.

5. Cibulskis RE, Bell D, Christophel EM, et al. Estimating trends in the burden of malaria at country level. Am J Trop Med Hyg 2007; 77(6 Suppl): 133–7.

6. van Eijk AM, Hill J, Noor AM, Snow RW, ter Kuile FO. Prevalence of malaria infection in pregnant women compared with children for tracking malaria transmission in sub-Saharan Africa: a systematic review and meta-analysis. Lancet Glob Health 2015; 3(10): e617–28.

7. Willilo RA, Molteni F, Mandike R, et al. Pregnant women and infants as sentinel populations to monitor prevalence of malaria: results of pilot study in Lake Zone of Tanzania. Malar J 2016; 15(1): 392.

8. Hellewell J, Walker P, Ghani A, Rao B, Churcher TS. Using ante-natal clinic prevalence data to monitor temporal changes in malaria incidence in a humanitarian setting in the Democratic Republic of Congo. Malar J 2018; 17(1): 312.

9. Brunner NC, Chacky F, Mandike R, et al. The potential of pregnant women as a sentinel population for malaria surveillance. Malar J 2019; 18(1): 370.

10. Kitojo C, Gutman JR, Chacky F, et al. Estimating malaria burden among pregnant women using data from antenatal care centres in Tanzania: a population-based study. Lancet Glob Health 2019; 7(12): e1695–e705.

11. Mayor A, Menendez C, Walker PGT. Targeting Pregnant Women for Malaria Surveillance. Trends Parasitol 2019; 35(9): 677–86.

12. Brokhattingen N, Matambisso G, da Silva C, et al. Genomic malaria surveillance of antenatal care users detects reduced transmission following elimination interventions in Mozambique. Nat Commun 2024; 15(1): 2402.

13. Ferrari G, Ntuku HM, Schmidlin S, Diboulo E, Tshefu AK, Lengeler C. A malaria risk map of Kinshasa, Democratic Republic of Congo. Malar J 2016; 15: 27.

14. Fanello C, Lee SJ, Bancone G, et al. Prevalence and Risk Factors of Neonatal Hyperbilirubinemia in a Semi-Rural Area of the Democratic Republic of Congo: A Cohort Study. Am J Trop Med Hyg 2023; 109(4): 965–74.

15. Signorell A. DescTools: Tools for Descriptive Statistics. R package version 0.99.54. 2024.

16. Wood DE, Lu J, Langmead B. Improved metagenomic analysis with Kraken 2. Genome Biol 2019; 20(1): 257.

17. MalariaGen, Ahouidi A, Ali M, et al. An open dataset of Plasmodium falciparum genome variation in 7,000 worldwide samples. Wellcome Open Res 2021; 6: 42.

18. Ariey F, Witkowski B, Amaratunga C, et al. A molecular marker of artemisinin-resistant Plasmodium falciparum malaria. Nature 2014; 505(7481): 50–5.

19. WHO. Artemisinin resistance and artemisinin-based combination therapy efficacy: status report: WHO, 2018.

20. Miotto O, Amato R, Ashley EA, et al. Genetic architecture of artemisinin-resistant Plasmodium falciparum. Nat Genet 2015; 47(3): 226–34.

21. Naidoo I, Roper C. Mapping ‘partially resistant’, ‘fully resistant’, and ‘super resistant’ malaria. Trends Parasitol 2013; 29(10): 505–15.

22. Lynch C, Pearce R, Pota H, et al. Emergence of a dhfr mutation conferring high-level drug resistance in Plasmodium falciparum populations from southwest Uganda. J Infect Dis 2008; 197(11): 1598–604.

23. Fidock DA, Nomura T, Talley AK, et al. Mutations in the P. falciparum digestive vacuole transmembrane protein PfCRT and evidence for their role in chloroquine resistance. Mol Cell 2000; 6(4): 861–71.

24. Happi CT, Gbotosho GO, Folarin OA, et al. Association between mutations in Plasmodium falciparum chloroquine resistance transporter and P. falciparum multidrug resistance 1 genes and in vivo amodiaquine resistance in P. falciparum malaria-infected children in Nigeria. Am J Trop Med Hyg 2006; 75(1): 155–61.

25. Chang HH, Worby CJ, Yeka A, et al. THE REAL McCOIL: A method for the concurrent estimation of the complexity of infection and SNP allele frequency for malaria parasites. PLoS Comput Biol 2017; 13(1): e1005348.

26. Schneider KA, Tsoungui Obama HCJ, Kamanga G, Kayanula L, Adil Mahmoud Yousif N. The many definitions of multiplicity of infection. Front Epidemiol 2022; 2: 961593.

27. Mesia Kahunu G, Wellmann Thomsen S, Wellmann Thomsen L, et al. Identification of the PfK13 mutations R561H and P441L in the Democratic Republic of Congo. Int J Infect Dis 2024; 139: 41–9.

28. Mita T, Venkatesan M, Ohashi J, et al. Limited geographical origin and global spread of sulfadoxine-resistant dhps alleles in Plasmodium falciparum populations. J Infect Dis 2011; 204(12): 1980–8.

29. Plowe CV. Malaria chemoprevention and drug resistance: a review of the literature and policy implications. Malar J 2022; 21(1): 104.

30. Kajubi R, Ochieng T, Kakuru A, et al. Monthly sulfadoxine-pyrimethamine versus dihydroartemisinin-piperaquine for intermittent preventive treatment of malaria in pregnancy: a double-blind, randomised, controlled, superiority trial. Lancet 2019; 393(10179): 1428–39.

31. Sisowath C, Ferreira PE, Bustamante LY, et al. The role of pfmdr1 in Plasmodium falciparum tolerance to artemether-lumefantrine in Africa. Trop Med Int Health 2007; 12(6): 736–42.

